# An autoantibody-based machine learning classifier for the detection of early-stage non-small cell lung cancer

**DOI:** 10.1101/2025.01.09.24315050

**Authors:** Andrew F Kung, Chukwuka A Didigu, Chung-Yu Wang, Aditi Saxena, Bryan Castillo-Rojas, Anthea M Mitchell, Sabrina A Mann, Alyssa Murillo, Kelsey C Zorn, Lloyd Bod, David M Jablons, Johannes R Kratz, Joseph L DeRisi

**Author notes:** Correspondence should be addressed to Joseph DeRisi < >.

## Abstract

The humoral immune system plays a significant role in the immune response to cancer but is challenging to study at scale. We used programmable phage immunoprecipitation sequencing (PhIP-Seq) to profile the autoantibody repertoire in non-small cell lung cancer (NSCLC) patients for the purpose of training a machine learning-based classifier to distinguish NSCLC patients from healthy controls using 301 primarily early-stage, asymptomatic NSCLC patients and 352 healthy controls. The classifier performed well in cross-validation (average ROC-AUC = 0.94) and in an independently analyzed clinical validation cohort of 134 NSCLC patients and 96 healthy controls (ROC-AUC = 0.84). Classification performance can be maintained with only a few hundred target peptides, provided a sufficiently large cohort is used for optimal training. Our findings suggest the existence of a measurable autoreactive humoral profile in NSCLC and demonstrate the potential for serum-based early detection of cancer independent of nucleic acids.

## Introduction

Non-small cell lung cancer (NSCLC) is the leading cause of cancer deaths in the United States, accounting for an estimated 238,340 new cases and 127,070 deaths in 2023 ^1^. Although the prognosis of NSCLC improves significantly when diagnosed at an earlier stage (5-year survival of 61% at Stage I vs. 6% at Stage IV), up to half of NSCLC patients have stage IV disease at the time of diagnosis ^2^. While low-dose chest CT (LDCT) scans are effective in screening for lung cancer in patients with a significant smoking history ^3^, fewer than 5% of eligible patients undergo LDCT screening, with cost and convenience serving as barriers to widespread adoption ^4–6^. Additionally, the current LDCT recommendation does not extend to patients who have never smoked, even though the incidence of never-smoker lung cancer continues to increase in the United States, particularly in women ^7,8^, with these patients now accounting for up to one in four new NSCLC diagnoses ^9,10^. Widespread adoption of LDCT amongst broader groups presents its own challenges, given the rate and burden of false positives and incidental findings ^11,12^. These hazards would undoubtedly increase as prospective diagnostic screening methodologies such as LDCT continue to gain traction in lower-risk populations ^13^. These considerations, taken together, highlight the need for complementary novel screening approaches that can identify patients at-risk for NSCLC and be deployed at scale across the population. Typical liquid biopsy tests detect cancer by measuring analytes in the blood, such as circulating tumor DNA, RNA, and circulating tumor cells. Recent examples include whole-genome sequencing of cell-free DNA ^14,15^ and multi-cancer methylation markers ^16^. While these tests can identify actionable mutations in late-stage disease, issues of sensitivity have limited their use in the *de novo* detection of early-stage cancers ^17^.

The adaptive immune system plays a significant role in the earliest immune responses to cancer, from the removal of aberrant cells before they become cancerous (immunosurveillance) to the equilibrium-like maintenance of more advanced lesions (immunoediting) ^18^. The presence of an immune response to cancer at its earliest stages presents a unique opportunity to detect the presence of a tumor. Indeed, the understanding and co-opting of these mechanisms has been foundational to the field of cancer immunotherapies. Most scientific and clinical efforts have been focused on the T-cell (cell-mediated) response rather than the B-cell (antibody-mediated) response, despite evidence that both play important roles in the immune response to cancer. The majority of previous attempts at profiling the antibody response to cancer-specific antigens or self-antigens have relied on a candidate approach to build antibody panels for cancer screening and prognosis ^19–21^. While many of these approaches have shown early promise, validation on larger cohorts has faced challenges ^22^, with large-scale clinical utility yet to be determined.

Programmable phage display and phage immunoprecipitation sequencing (PhIP-Seq) is a powerful tool for comprehensively profiling the humoral immune repertoire with proteome-wide resolution ^23,24^. As opposed to classical phage display, wherein short random peptides are displayed, programmable phage display leverages large-scale oligonucleotide synthesis to create libraries of long (40+ amino acid) overlapping peptides spanning a given proteome of protein-coding sequences, encoded and displayed by T7 bacteriophage. These phage libraries are applied to iterative cycles of immunoprecipitation pull-downs with patient immunoglobins, followed by next-generation sequencing to quantify individual peptide enrichments relative to the pre-selected library, thereby generating an antibody “autoreactome” signature from each patient sample ^25^. PhIP-Seq has successfully identified autoimmune targets associated with a wide range of diseases, including, but not limited to, genetic immune dysregulation ^26^, paraneoplastic diseases ^24,27^, rare childhood disorders (Mandel-Brehm et al., 2022), and cancer immunotherapy-related adverse events ^28^. Outside of cancer associated with paraneoplastic disease, however, this technique has not yet been applied to identify a broader antibody signature associated with cancer for detection and classification purposes.

Several PhIP-Seq analysis approaches have been described, generally relying on various statistical techniques to determine a set of differentially enriched peptides or proteins ^24,26,29–31^. Basic machine learning approaches have been previously applied to PhIP-Seq data ^32^, classifying patients with a known monogenic autoimmune disorder, autoimmune polyendocrine syndrome type 1 (APS-1), versus healthy controls solely based on PhIP-Seq signatures. These models identified previously known and novel antigens that were subsequently validated through orthogonal assays. While diseases such as APS1 represent dramatic shifts in the immune repertoire, early cancers may present comparatively fewer and perhaps more subtle changes. Therefore, more advanced machine learning models may be necessary to interrogate such phenotypes thoroughly. Indeed, machine learning, particularly deep learning, has been successfully applied in other contexts of cancer immunology, including HLA-specific neoantigen mass-spectrometry ^33^ and T-cell receptor sequencing ^34^. Machine learning with VirScan, PhIP-Seq on viral epitopes, has been used to highlight at-risk patients for hepatocellular carcinoma through a signature primarily driven by hepatitis C infection ^35^. Here, we applied and evaluated machine learning techniques to derive cancer-specific signatures following PhIP-Seq profiling of plasma from patients with non-small cell lung cancer (NSCLC) as well as healthy individuals, developing a highly predictive classifier of NSCLC. Our results suggest a novel and robust alternative to existing nucleic acid-based liquid biopsy techniques for early cancer detection.

## Results

### Classification of NSCLC versus healthy control

To ascertain the potential of autoantibody profiling for cancer classification, we performed PhIP-Seq immunological profiling on a cohort of plasma samples previously collected from 301 patients with NSCLC at the University of California, San Francisco (UCSF) Thoracic Oncology Laboratory and plasma from 352 healthy volunteers (**Table 1, Figure 1**). An important feature of this NSCLC cohort is its early-stage composition, with more than 90 percent of patients having Stage I or II disease. Similarly, over two-thirds of patients in this training cohort were asymptomatic at the time of diagnosis.

**Figure 1:**
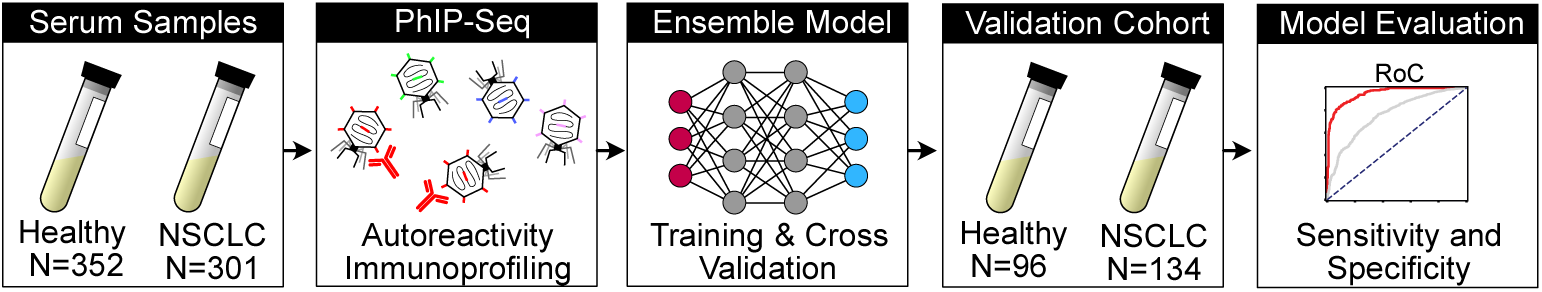
Schematic of PhIP-Seq signature-based NSCLC classification. Workflow of a potential cancer diagnostic pipeline based on humoral signature. A patient’s serum is run on the PhIP-Seq assay, and the results are fed into our bagged machine learning model (chosen after evaluation with 10-fold cross-validation), which outputs a probability of tumor presence. Performance was maintained when testing on an independent validation cohort.

**Table 1.** Summary of Patient Clinical and Pathological Characteristics. Table 1: Cohort information Clinical cohort demographics of the primary and validation cohorts of non-small cell lung cancer patients from UCSF Thoracic Oncology, as well as available information from the cohorts of healthy controls.

|  | Training Cohort | Validation Cohort |
| --- | --- | --- |
| **NSCLC Patients** | **301** | **134** |
| Age | 68.7 (9.8)<sup>§</sup> | 68.8 (10.3)<sup>§</sup> |
| Females | 189 (62.8) | 81 (60.4) |
| Males | 112 (38.2) | 53 (39.6) |
| Smoking History |  |  |
| Yes | 191 (63.5) | 80 (58.4) |
| No | 110 (36.5) | 57 (41.6) |
| Symptomatic |  |  |
| Yes | 101 (33.6) | 54 (40.3) |
| No | 200 (66.4) | 80 (59.7) |
| Stage |  |  |
| 0 | 1 (0.3) | 3 (2.2) |
| I | 231 (76.7) | 70 (52.2) |
| IA1 | 67 (22.3) | 12 (9.0) |
| IA2 | 67 (22.3) | 22 (16.4) |
| IA3 | 22 (7.3) | 16 (11.9) |
| IB | 75 (24.9) | 19 (14.2) |
| II | 43 (14.3) | 38 (28.4) |
| IIA | 12 (4.0) | 9 (6.7) |
| IIB | 31 (10.3) | 29 (21.6) |
| III | 25 (8.3) | 20 (14.9) |
| IIIA | 23 (7.6) | 15 (11.2) |
| IIIB | 2 (0.7) | 5 (3.7) |
| IIIC | 0 (0.0) | 0 |
| IV | 1 (0.3) | 2 (1.5) |
| **Healthy Controls** | **352** | **96** |
| Age | 38.7 (12.6)<sup>§</sup> | 29.3 (9.0)<sup>§</sup> |
| Females | 176 (50.0) | 54 (56.2) |
| Males | 176 (50.0) | 42 (45.8) |

Numbers in parentheses represent the percentage of patients except where stated.
<sup>§</sup>Cohort Mean (Standard Deviation)

Following our previously described high-throughput PhIP-Seq protocol ^32,36^, autoreactive serological profiles of the entire cohort were captured. A detailed protocol is available at protocols.io (<http://dx.doi.org/10.17504/protocols.io.ewov1459kvr2/v>). Briefly, plasma from patients and controls were randomly arrayed in 96-well plates, then incubated with a previously validated custom library of 730,000+ unique phage species spanning the entire annotated “healthy” human proteome, including known isoforms, tiled across 49 amino acid fragments with a 25 amino acid overlap ^24^. Following three rounds of enrichment through immunoprecipitation of patient or healthy control antibodies bound by protein A/G beads, the resultant enriched phage species were sequenced using the Illumina NovaSeq platform (median read depth of 1,276,108 reads per sample) and aligned to the reference with RAPSearch2 ^37^. To ensure consistency and to check for potential batch effects, a subset of the samples was run in duplicate on separate plates. Duplicates on separate plates demonstrated a sample-wise correlation (average Pearson coefficient > 0.80) to its corresponding replicate as well as to a separate PhIP-Seq run on an earlier date with limited correlation to other samples in the set (**Supplemental Figure 1**). The complete PhIP-Seq data is available on Dryad at <https://doi.org/10.5061/dryad.08kprr5bk>.

Enriched peptide counts, scaled for sequencing depth to reads-per-100,000 (RP100K), were used as input into a range of machine learning classifier models, both on the individual peptide level (731,724 unique features), as well as collapsed by associated protein to account for tiling and isoform redundancy (19,998 unique features), similar to previous analyses with the same library ^26^. Classifiers, described below, were initially evaluated using 10-fold cross-validation - systematically withholding 10% of the samples (both case or control), training on the remainder of the set, and then predicting the status of the withheld samples (**Figure 1**).

A range of machine learning classifiers, implemented using the popular *scikit-learn* and *PyTorch* python packages, were evaluated, including logistic regression, neural networks, support vector machines, and random forests. At a range of specificity thresholds, logistic regression and neural networks consistently outperformed random forest, both for peptides and proteins (**Figure 2A**). After hyperparameter optimization on individual levels, we found that an ensemble model incorporating predictions from multiple models yielded the best classification performance, with an average area under the receiver operator curve (AUC-ROC) of 0.94 after 1000 bootstrapped iterations (**Figure 2B**). Ensemble models have previously been successfully implemented in other biological classification tasks ^38–41^. The three components of the ensemble model were logistic regression on the peptide level, logistic regression on the protein level, and a fully connected feed-forward neural network on the peptide level (see Methods and GitHub repository <github.com/afkung/nsclc-classify> for model details and specific packages used). Although not necessarily optimal, an ensemble model provided the biological comprehensibility of the linear components and latent benefits captured by the non-linear components. Indeed, performance was superior to that of the individual models in isolation, with a sensitivity of 70% at a specificity of 95% (Supplemental Table 1). Given this proof-of-concept, it is likely that more sophisticated models have the potential to further increase performance.

**Figure 2:**
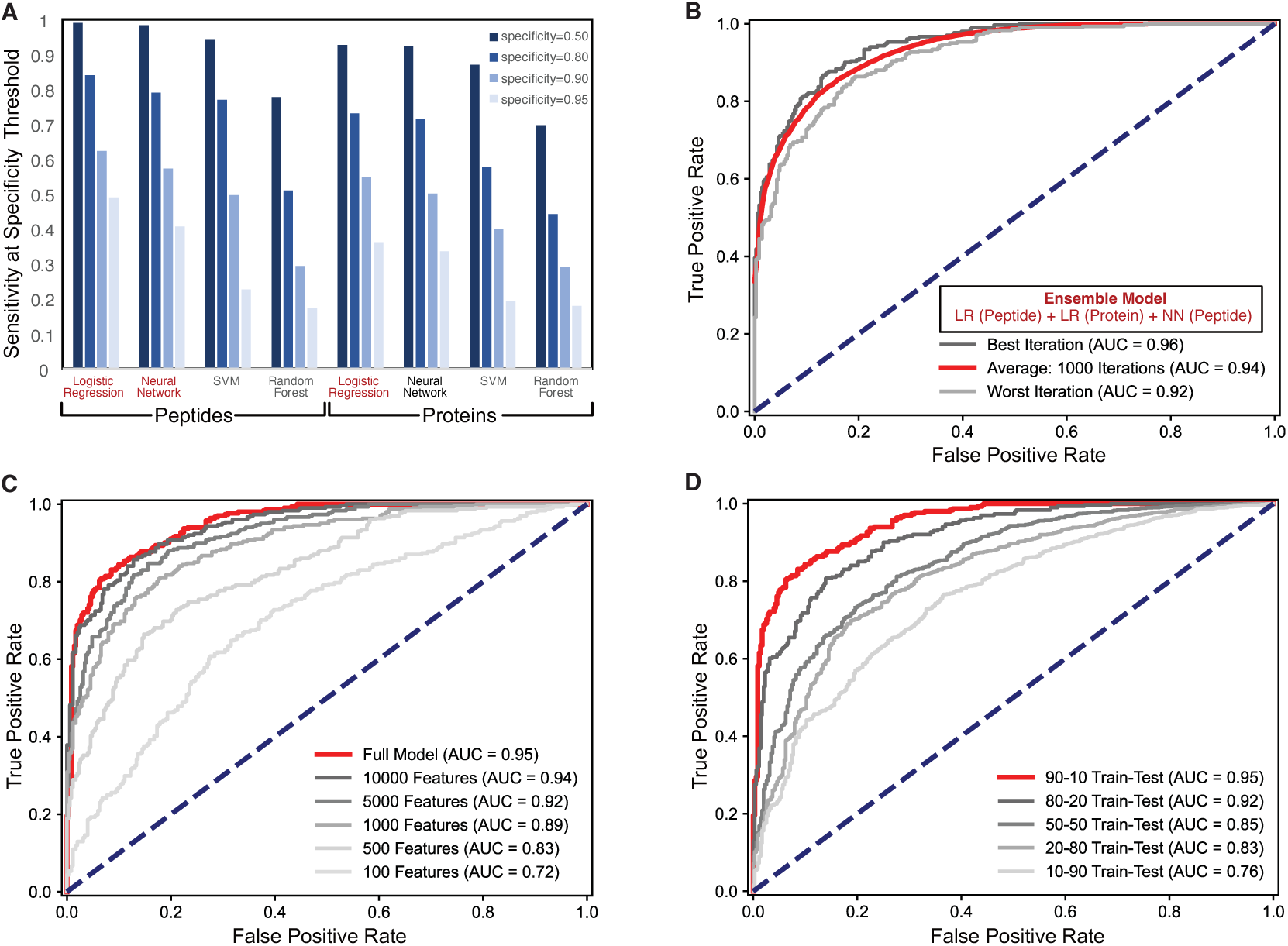
Classifier performance and parameterization. A. Comparison of individual component model sensitivity at various specificity thresholds (0.50, 0.80, 0.90, 0.95) for various models on both gene-level and peptide level, with asterisks denoting component models. B. Receiver-operator-characteristic (ROC) curve for the classification of NSCLC vs. healthy controls based on a bagged model, the best performing classifier (bootstrapped across 1000 iterations, with an average ROC-AUC = 0.94). C. ROC curves of varying feature space sizes, determined by maximum RP100K threshold across all samples, NSCLC and healthy control. D. ROC curves of varying train-test split sizes, across NSCLC and healthy control, cross-validated.

### Model parameterization suggests a robust and specific signal

The best-performing ensemble model was further characterized through parameterization. While the full model utilized all values in the dataset, we evaluated the model training and cross-validation performance by progressively limiting the number of peptide-level input features from 100 to 100,000 features relative to the complete dataset. (**Figure 2C**). As expected for zero-inflated data structures like PhIP-Seq peptide/protein counts, removing low-level and zero counts did not affect the model’s performance. Indeed, removing more than 99% of available features (leaving 5,000 features remaining) still yielded a comparable AUC (0.92 average). However, model performance suffered progressively more degradation when restricting input features to 1,000 or less (∼0.1% of the possible features). At 100 features, the average AUC had decreased to 0.72. This suggests that the underlying information content separating case and healthy control is distributed across a plurality of autoreactive protein targets. This is in contrast to cancer-related autoimmune disorders, such as seminoma-associated paraneoplastic encephalitis, where a single autoreactive target (KLHL11) is sufficiently diagnostic ^27^.

To further characterize the specificity of the classifier, the train-test split sizes of the model were systematically varied, training on 90%, 80%, 50%, 20%, and 10% of the samples (both NSCLC and healthy, cross-validated) and testing on the remainder (**Figure 2D**). As expected, model performance progressively degraded as the training split size decreased. However, the model maintained reasonable classification ability (AUC > 0.80) with as little as 20% of the samples (**Figure 2C**). These data suggest the presence of a shared humoral autoreactive signal across multiple individuals among cases and not controls while simultaneously reinforcing the need for sufficiently large cohorts for optimal training.

### Top features identified by model are orthogonally validated and indicate potential tissue and cancer specificity

Although the logistic regression model alone did not have the best classification performance among the models tested here, it has the distinct advantage of being a linear model that can extract and quantify the contributions of the input features. Four peptides associated with top coefficients from the model (PRRC2A, QTRT1, NRAC, LAX1) were chosen for separate experimental validation via a previously described split-luciferase binding assay (SLBA) ^42^. Overall, plasma from NSCLC patients yielded significantly higher binding to peptides encoded by these four genes than plasma from non-cancer controls (**Figure 3A**), although there was a wide range of values reinforcing the notion that a small number of features would likely have inadequate predictive value alone. Similarly, RNA expression profiles from paired cancer-normal samples within The Cancer Genome Atlas (TCGA-LUAD) for these genes (PRRC2A, QTRT1, and LAX1; NRAC was not available) all indicate a statistically significant increase in expression of these genes in the tumor compared to the paired normal lung tissue from the same patients (**Figure 2B**). When mapped to normal gene expression in the genotype-tissue expression atlas (GTEx), the Z-scores across all available tissue types of the top 25 features (NRAC not available) suggest lung-specific enrichment compared to other relevant tissue types (**Figure 3C**). When considering the average Z-score in broader sets of top features (25 to 100+ genes) the enrichment is inversely proportional to the number of features, suggesting that most tissue-specific signal resides in the top ranked coefficients.

**Figure 3:**
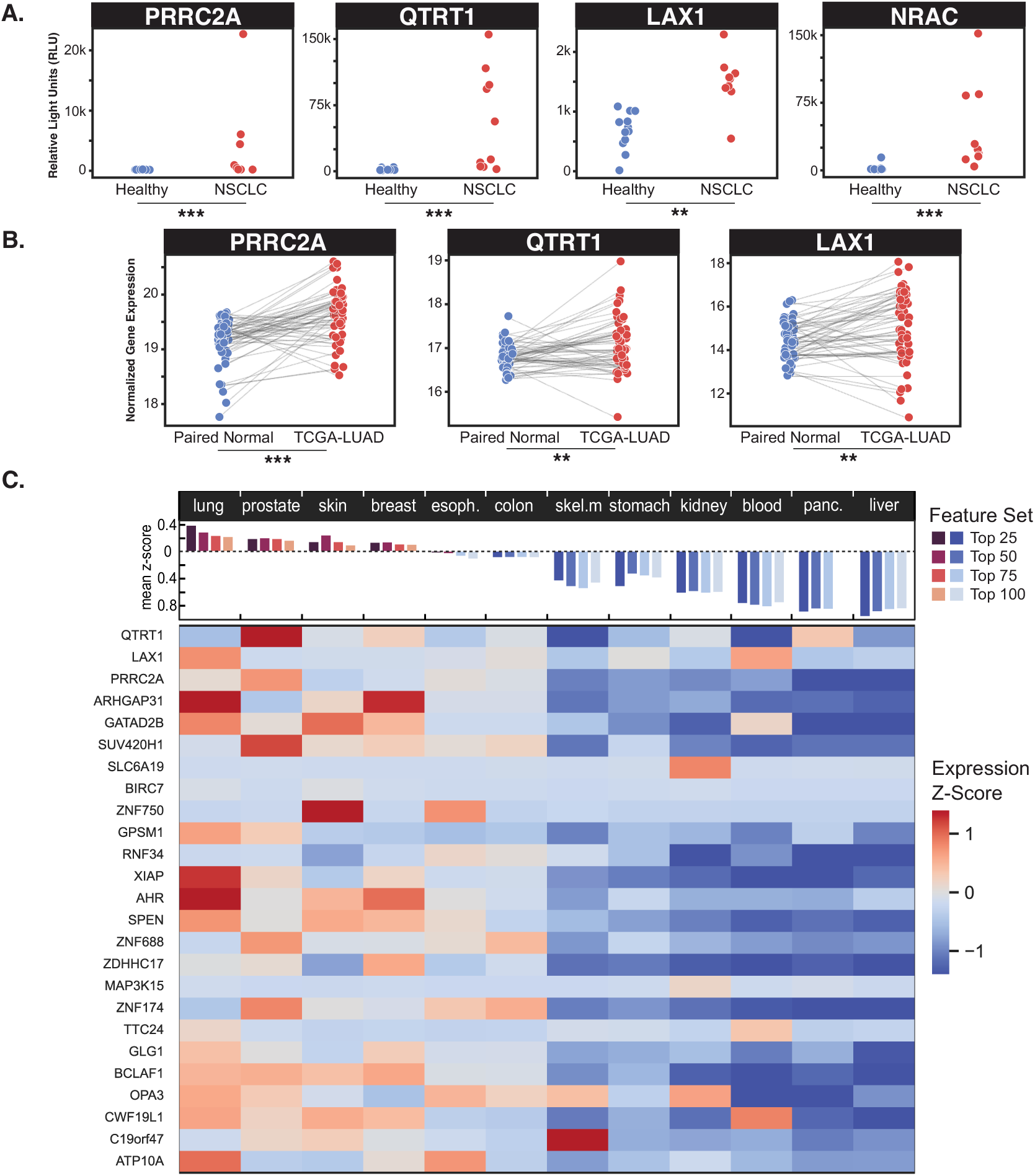
Interrogation of top features identified by linear models. A. Split-luciferase binding assay of four top features identified by the model’s linear components (PRRC2A, QTRT1, LAX1, NRAC) shows significantly enriched binding in NSCLC samples compared to healthy controls (Mann-Whitney U test: *** p = 0.0005, **** p < 0.0001). B. Three of the identified features have significantly enriched gene expression in The Cancer Genome Atlas for lung adenocarcinoma (TCGA-LUAD) compared to healthy lung tissue from the same patient (Wilcoxon signed-rank test: ** p < 0.002, *** p < 1e-5). NRAC was not available in TCGA. C. Gene expression in various healthy tissues from the Genotype-Tissue Expression (GTEx) indicates potential lung-specificity, when comparing Z-score across all available tissue types in the top 25 features, as well as the average of top feature sets (25, 50, 75, and 100) compared to relevant comparative tissue types.

### Independent cohort validation

Overfitting is a major concern for classification tasks. To test the robustness of the cancer signature, the model was trained with the entire discovery cohort (301 NSCLC samples and 352 healthy samples) and then subsequently used to evaluate a blinded cohort consisting of 134 NSCLC and 96 healthy samples (**Table 1**), all previously unseen by the model. Samples were processed in the same manner as before, and sequencing data was then transmitted, completely blinded with no pre-processing or evaluation, to an independent validator from an outside institution well-versed in PhIP-Seq data and analysis but otherwise previously uninvolved in the project. Using a web server, the independent validator could submit anonymous files containing peptide counts from the blinded samples for classification by our model. Our validator reported robust classification (ROC-AUC = 0.84, **Figure 4A**) and predictive value at various prespecified classifier thresholds in our independent validation cohort (**Figure 4B**). As expected, the performance of the model on this independent cohort was less than the training cohort. Various test manipulations performed by the independent validator produced predictable results from the model that were in line with expectations, underlying the validity of the independent evaluation. Shuffling samples and count orders had no effect. Introducing a uniform background distribution by adding 50 to all counts did not significantly alter the result, while addition of random peptide values completely ablated the signal (**Figure 4C**). This blinded independent third-party evaluation on separate samples, never used in training, suggests the model is reproducible for early-stage cancer.

**Figure 4:**
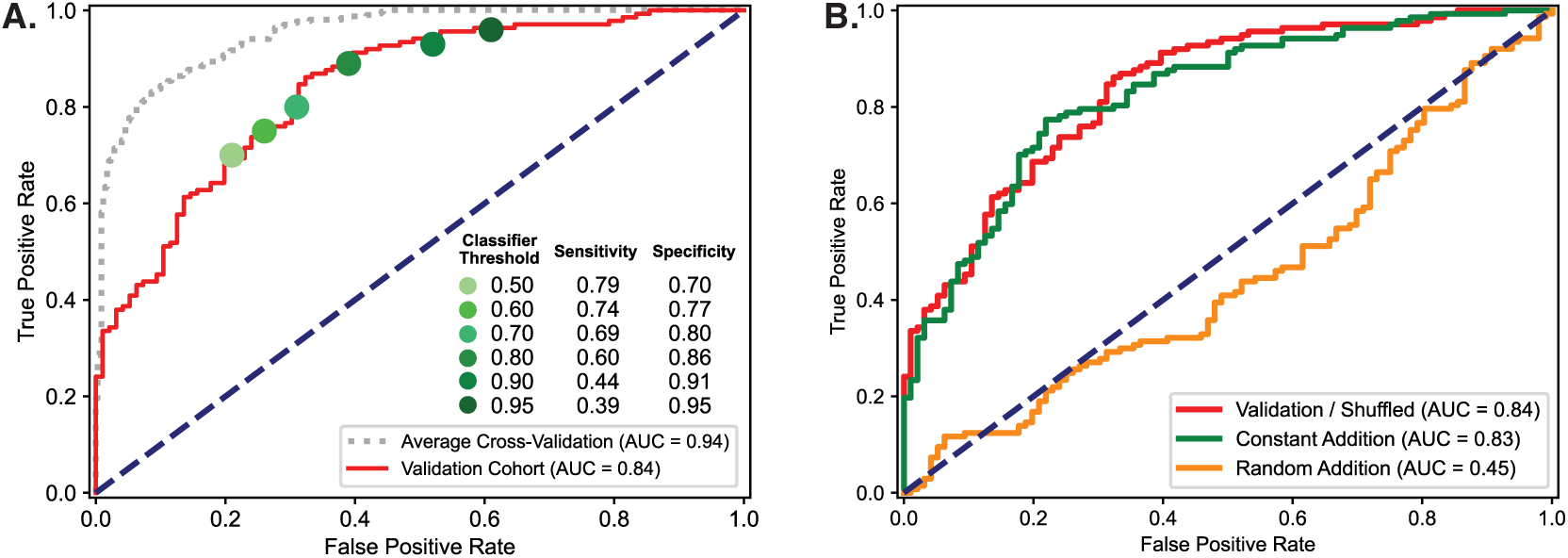
Validating classifier performance with independent NSCLC cohort. A. ROC curve of classification of independent blinded validation cohort (137 NSCLC and 96 healthy, all previously unseen) with model trained on entire initial discovery cohort (301 NSCLC and 352 healthy). Predictive power decreased compared to cross-validation, as expected, but remained quite robust (ROC-AUC = 0.84). B. Sensitivity and specificity for validation cohort at various classification thresholds. C. Manipulated input testing performed as expected - no effect with input re-ordering (ROC-AUC = 0.84), a small decrease when a constant input of 50 reads was systematically added to all samples (ROC-AUC = 0.83), and complete signal ablation with random inputs (ROC-AUC = 0.44).

### Correlation to relevant clinical features

Certain clinical features in NSCLC are linked to disease development, progression, and outcome. When sub-setting the training cohort and validation cohorts by these features, the distribution of prediction values from the classifier (0 as healthy, 1 as NSCLC) offers insights into the captured signal. Consistent with the ROC curves, there were robust and significant differences between NSCLC and healthy for both the cross-validated training and independent clinical validation cohorts (**Figure 5A**). Tobacco use is a well-established risk factor for NSCLC, and smoking history (self-reported current or former) was indeed correlated with higher prediction scores, significantly so in the validation cohort (0.678 vs. 0.799, p=0.011) (**Figure 5B**). Tumor stage represents the size and invasiveness of the disease, with later stages representing a larger global tumor footprint. Interestingly, we did not observe significant differences in classifier performance in patients with early versus late-stage disease in either cohort (**Figure 5C**). This starkly contrasts with most nucleic acid-based early-detection modalities, which typically see an absence of signal in earlier-stage disease due to the naturally lower amount of circulating tumor DNA (ctDNA) present in early-stage disease.

**Figure 5:**
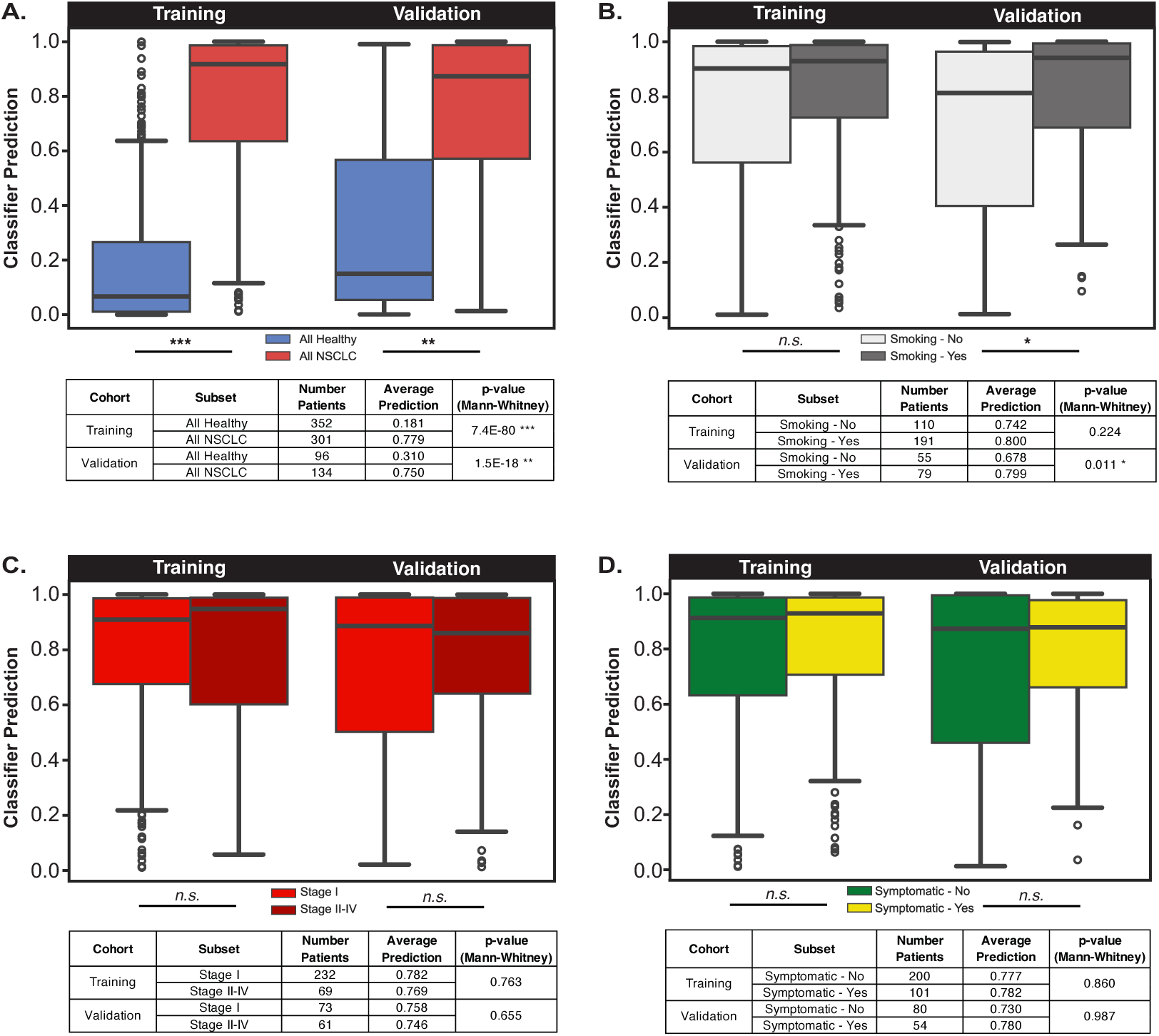
Subsetting cohort by relevant clinical features. A. Distributions of classifier predictions (0 representing healthy, 1 representing NSCLC) for all samples in both training and validation cohorts. Significant differences were seen in both cohorts (Mann-Whitney U-test). B. Distributions of classifier predictions split by smoking status. Predictions for smokers were consistently higher, significantly so in the validation cohort. C. Distributions of classifier predictions split by cancer stage (Stage I vs. Stages II-IV). No significant difference was observed. D. Distributions of classifier predictions split by symptomatic status at time of diagnosis. As in 5C, no significant differences were observed.

Similarly, there were no significant differences in classifier performance among patients who were symptomatic (defined as any of the following: chest pain, cough, hemoptysis, dyspnea, fatigue, weight loss) versus asymptomatic patients at the time of diagnosis in either the training or validation cohorts (**Figure 5D**).

## Discussion

We used PhIP-Seq to systematically profile the proteome-wide humoral response to NSCLC with the specific intent of testing whether autoreactome profiles could be used as the basis for a machine learning-based classifier for early-stage lung cancer. We find that these autoreactive profiles distinguish NSCLC patients from healthy controls with favorable performance characteristics. Furthermore, classifier performance was maintained in a blinded independent clinical validation cohort analyzed at a separate institution, thus highlighting the robustness of this approach.

Overexpression and tissue-aberrant expression of normal proteins is a known hallmark of cancer. In support of this, analysis of top autoantibody targets driving our classifier revealed that these proteins are preferentially enriched in tumor tissue when compared to normal lung tissue within the same patients, and may have greater lung-specific expression compared to other tissue types. Increased expression of the genes corresponding to the validated peptides have been previously implicated in poor prognosis in lung cancer for QTRT1 ^43^ and immune infiltration for PRRC2A ^44^. The lung-specific nature of the autoantibody targets driving the classifier underscores the biological significance of this approach and suggests it can be successfully applied to other tumor types, with expanded tumor-specific training sets.

A known issue among nucleic acid-based early detection approaches for cancer is a significant decrease in test performance in early-stage and asymptomatic patients when compared to late-stage, symptomatic patients ^45–47^. In contrast, our classifier performance was not significantly different in patients with Stage I disease when compared to patients with Stage II-IV disease and in patients who were asymptomatic at the time of sample collection. Interrogating the humoral response has the advantage of leveraging the natural signal amplification that occurs during the immune response to cancer cells, unlike technologies that test for cancer-cell derived molecules, which will always be dependent on the quantity of those cells present. Coupled with iterative rounds of enrichment using PhIP-Seq, autoantibody profiling offers a unique way to capitalize on the immune response to cancer, a response present in the earliest stages, and thus a means to identify patients with occult cancers. This has important implications for future cancer screening modalities, as earlier diagnosis is associated with markedly improved outcomes.

Using conventional machine learning techniques, this study is a proof-of-concept for cancer detection via analysis of PhIP-Seq data. Moving from a hit-calling analytical paradigm to a signature-based one offers tremendous promise for diagnostic serological profiling beyond the classification tasks shown here. Whereas a single linear model on the protein level was sufficient for the classification of APS1 versus healthy controls ^32^, the improved performance of an ensemble model indicates that the component models provide complementary and nonredundant information. Given the tiling and isoform redundancy built into our peptide library ^24^, collapsing by protein may boost the signal from lower-abundance single peptides with shared sequences (both exact matches and biochemical similarities). Similarly, neural networks may recognize meaningful interactions between seemingly unrelated peptides, potentially indicative of amino acid sequence similarity and biological relationships (such as shared pathways) undetected by linear models. Although the model type and hyperparameter spaces were not fully explored in this study, the robust performance of an ensemble model with relatively standard machine learning components serves as an encouraging proof-of-concept.

The use of linear model components allowed the extraction of features that were experimentally validated and biologically meaningful, while the non-linear components appear to add latent value. More advanced models, such as attention-based neural networks or multi-modal models incorporating features beyond numerical autoantibody enrichment, have been successfully applied to other biological contexts and may improve classification performance ^48–51^.

This study has several limitations. The study was performed with a peptide library spanning the entire human proteome and, therefore, does not contain sequences encoded by tumor-specific neoantigens. Autoantibodies to neoantigens have been previously described ^52^ and represent promising theoretical targets but would likely not be detected in this assay. Furthermore, the phage-encoded peptide library in this work cannot display protein folds that exceed 47 amino acids or post-translational modifications, such as glycosylation. Humoral responses to purely conformational or modified epitopes, which have been previously characterized ^53^, are unlikely to contribute to the signatures detected here. Incorporation of additional cancer-specific features has the potential to further improve training and model performance.

Another limitation of this study is the sourcing of NSCLC samples and available healthy controls. As the NSCLC samples were collected from a single center, future studies are needed to ensure the classifier is generalizable to a broader population of NSCLC patients. The healthy controls used in this study were collected from numerous sources (New York Blood Center, UCSF community blood drive, SeraCare, BioIVT) raising the possibility of batch effects at the time of blood collection contributing to the performance of the classifier. Finally, limited clinical information was available for the healthy control samples beyond basic demographics (age, sex, race). Clinical characteristics with known cancer associations (such as smoking status, familial history, and long-term health outcomes) were unavailable for the presumed healthy controls. Therefore, we are not able to ascertain cancer risk factors or the development of future cancers within the healthy control cohort. The mean ages of the healthy and NSCLC samples used here were also different (38.7 vs. 68.7 for the training cohort). Although separation between NSCLC and healthy remained robust, we observed lower prediction values in younger (<50 years) NSCLC patients and higher prediction values in older (>50 years) healthy individuals (**Supplemental Figure 2**), potentially related to the accumulation of immunosurveillance-associated autoantibodies, latent disease, or smaller sample sizes. Future age-matched training sets with long-term outcomes data will be required to further disambiguate the contributions of age and cancer.

Taken together, these proof-of-concept results strongly suggest the presence of a distinct humoral autoreactive signal in patients with NSCLC that can be used to distinguish them from healthy controls using machine learning applied to PhIP-Seq autoantibody profiles. The ability to detect tumors using a serological autoantibody-based signature before the development of overt disease holds tremendous promise for improving the early detection of cancer and may be highly complementary to existing liquid-biopsy approaches.

## Materials and Methods

### Sample acquisition

The primary (n = 301) and validation (n = 134) cohorts of primarily early-stage NSCLC plasma samples were collected from the UCSF Thoracic Oncology Program (San Francisco, CA). All patients gave informed consent for sample collection. Samples were collected in accordance with UCSF ethical guidelines and regulations for the conduct of responsible research. The study was approved by the UCSF Institutional Review Board (IRB# 11-06107). Deidentified healthy control plasma was collected from four sources: courtesy of New York Blood Center (New York, NY), purchased from SeraCare (Milford, MA), purchased from BioIVT (Westbury, NY), and donors from a UCSF community drive (UCSF IRB# 22-3611; San Francisco, CA).

### Phage immunoprecipitation sequencing (PhIP-Seq) assay

PhIP-Seq was run on the vacuum-based high-throughput assay described in Vazquez et al., 2022. Full protocol available here: <http://dx.doi.org/10.17504/protocols.io.ewov1459kvr2/v> Briefly, plasma samples across all patient cohorts were randomly arrayed across 96-well plates. In each well, 1 μL of patient plasma was incubated with 500 μL of our custom T7 phage display library of 731,724 unique species spanning the “normal” human proteome from NCBI RefSeq (Agilent Technologies, previously described in O’Donovan et al., 2020). After overnight incubation, antibodies were pulled down with Protein A/G beads (DynaMax). After three washes, the remaining bound-phage were then inoculated into a bacteria culture (BLT5403) and allowed to lyse. The process was then repeated with the resultant phage culture and fresh patient plasma twice more for a total of three iterative rounds. The final phage DNA was extracted and sequenced on the NovaSeq platform (Illumina).

### PhIP-Seq data analysis

Raw sequence reads were aligned and processed as previously described ^32,36^. Analysis was performed in Python with the following packages: *pandas, numpy*, *scipy, PyTorch*, and *scikit-learn.* Figures were made with *matplotlib*, *seaborn*, Adobe Illustrator, Affinity Designer, and Morpheus <https://software.broadinstitute.org/ morpheus>.

### Machine learning implementation and evaluation

Various machine learning models were implemented in Python with *scikit-learn* and *PyTorch*, and then evaluated through 10-fold cross-validation on the training set (352 NSCLC, 301 healthy), varying model parameters to maximize classification ability using ROC-AUC (*scikit-learn)* as the output metric. The following models and parameters were evaluated: Logistic regression (regularization, solver, iterations), Random forests (number of trees, depth, criterion, iterations), Support vector machine (regularization, kernel, iterations), and Neural network (activation function, loss function, number of hidden layers, layer size, iterations). The final ensemble model consists of logistic regression (L_2_ regularization, *liblinear* solver) on peptide and protein level and a fully-connected feed-forward neural network (*RELU* activation function on 2 hidden layers of sizes 100 and 50). The model details and implementation are available on github: <github.com/afkung/nsclc-classify>

### Split luciferase binding assay (SLBA)

SLBA was run as described in Rackaityte et al., 2023. Full protocol available here: <https://doi.org/10.17504/protocols.io.4r3l27b9pg1y/v1>

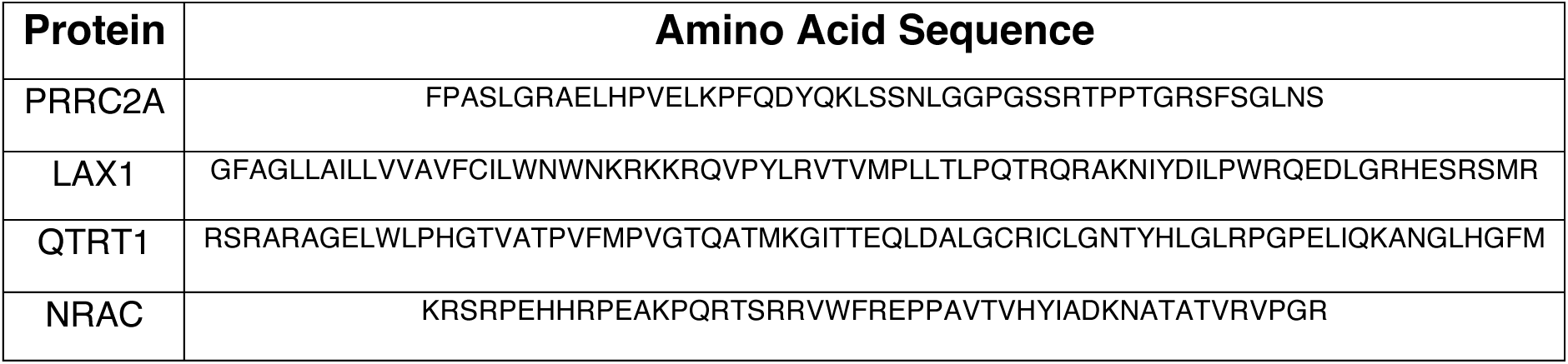

## Data Availability

All PhIP-Seq data is available on Dryad at < https://doi.org/10.5061/dryad.08kprr5bk>. Code associated with analysis is available on github <https://github.com/afkung/nsclc-classify>. Normal expression data from the Genotype-Tissue Expression Project (GTEx) used in the analyses described were downloaded from the GTEx Portal and dbGaP accession number phs000424.vN.pN. Lung adenocarcinoma data from The Cancer Genome Atlas Program (TCGA-LUAD) used in the analyses described in this manuscript were downloaded through the UCSC Xena Data Portal (University of California, Santa Cruz).

## Data Availability

All PhIP-Seq data is available on Dryad at ˂https://doi.org/10.5061/dryad.08kprr5bk˃. Code associated with analysis is available on github at ˂https://github.com/afkung/nsclc-classify˃. Normal expression data from the Genotype-Tissue Expression Project (GTEx) used in the analyses described were downloaded from the GTEx Portal and dbGaP accession number phs000424.vN.pN. Lung adenocarcinoma data from The Cancer Genome Atlas Program (TCGA-LUAD) used in the analyses described in this manuscript were downloaded through the UCSC Xena Data Portal (University of California, Santa Cruz).

https://doi.org/10.5061/dryad.08kprr5bk

https://github.com/afkung/nsclc-classify

## Acknowledgements

We would like to thank the New York Blood Center, Avail Bio, and members of the UCSF community for providing healthy control plasma samples. We would like to thank Hani Goodarzi, Jill Hollenbach, Yasin Senbabaoglu, and members of the DeRisi lab for useful comments and discussions. Funding was provided by the Chan Zuckerberg Biohub (San Francisco). L.B. was supported by grants from the Lung Cancer Research Foundation (LCRF) and the MGH Transformative Scholars Program.

---

**Supplemental Figure 1:**
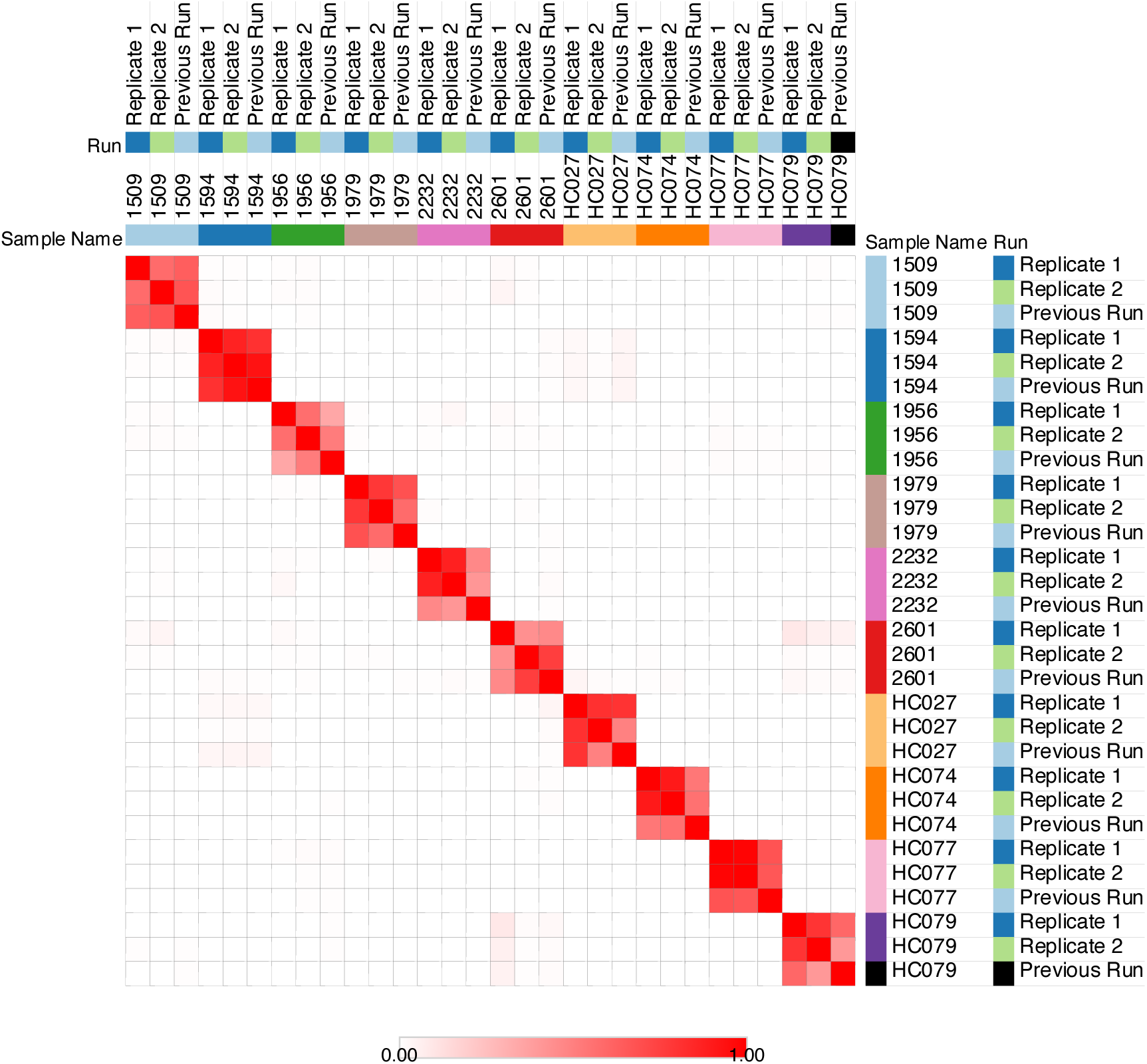
stability / reproducibility of signal Hierarchical clustering of Pearson correlations between PhIP-Seq samples shows that the autoantibody “signature” is consistent across replicates from a single run (in separate 96-well plates) as well as across previous runs on different dates.

**Supplemental Figure 2:**
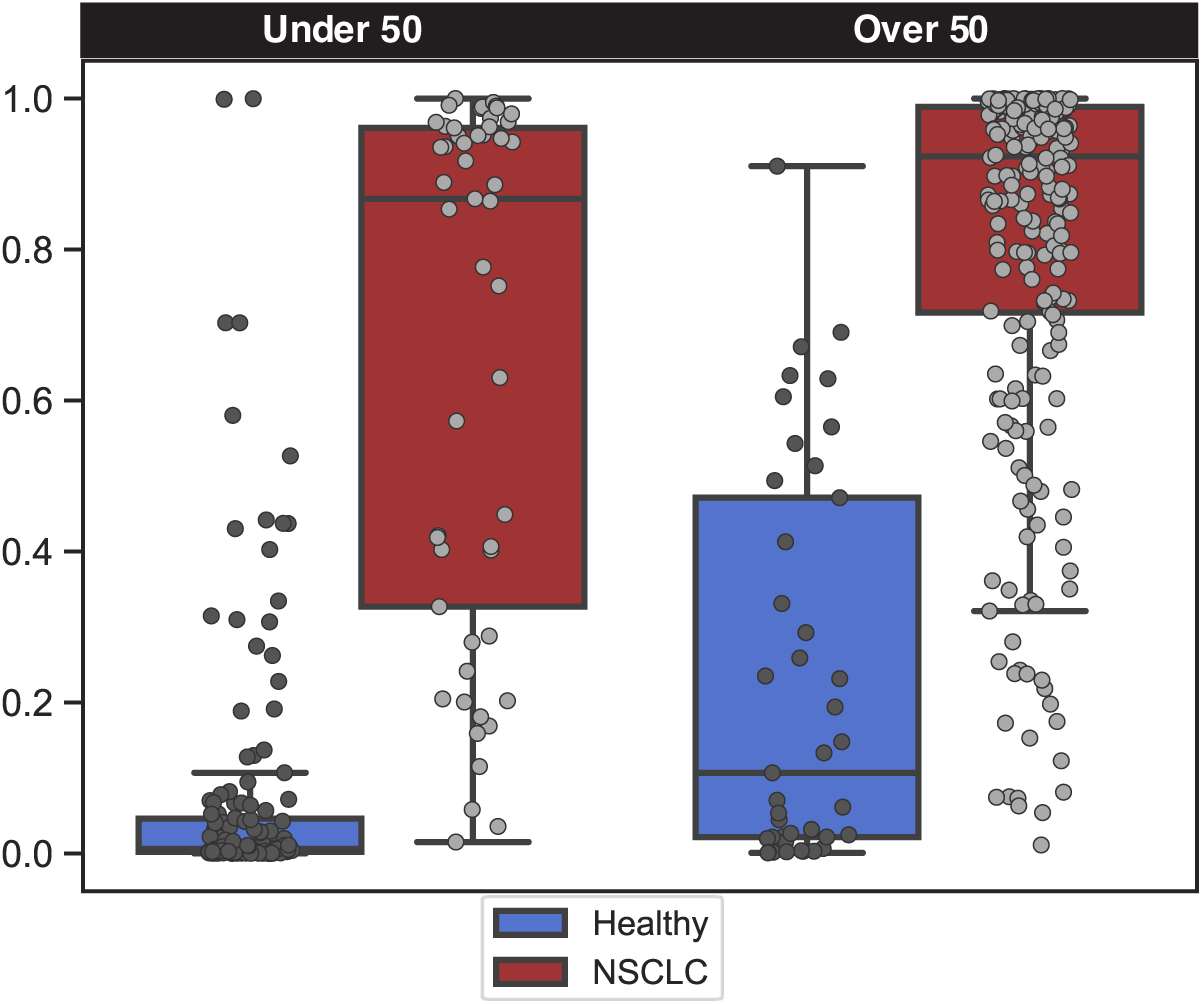
classification ability by age Prediction score of samples divided by age (under or over 50 years old). Although predictive power is worse for younger NSCLC patients and older healthy controls, potentially due to immunosurveillance theory or latent neoplasms, general separation remains robust in both groups.

## References

1. American Cancer Society. Cancer Facts & Figures 2023. (2023).

2. Siegel, R. L., Miller, K. D., Wagle, N. S. & Jemal, A. Cancer statistics, 2023. CA A Cancer J Clinicians 73, 17–48 (2023).

3. Mazzone, P. J. et al. Screening for Lung Cancer. Chest 160, e427–e494 (2021).

4. Jonas, D. E. et al. Screening for Lung Cancer With Low-Dose Computed Tomography: Updated Evidence Report and Systematic Review for the US Preventive Services Task Force. JAMA 325, 971 (2021).

5. Meza, R. et al. Evaluation of the Benefits and Harms of Lung Cancer Screening With Low-Dose Computed Tomography: Modeling Study for the US Preventive Services Task Force. JAMA 325, 988 (2021).

6. de Koning, H. J. et al. Reduced Lung-Cancer Mortality with Volume CT Screening in a Randomized Trial. N Engl J Med 382, 503–513 (2020).

7. Pelosof, L., et al. Proportion of Never-Smoker Non–Small Cell Lung Cancer Patients at Three Diverse Institutions. JNCI J Natl Cancer Inst 109, djw295 (2017).

8. Jemal, A. et al. The Burden of Lung Cancer in Women Compared With Men in the US. JAMA Oncol (2023) doi:10.1001/jamaoncol.2023.4415.

9. Cho, J. et al. Proportion and clinical features of never-smokers with non-small cell lung cancer. Chin J Cancer 36, 20 (2017).

10. Zhang, T. et al. Genomic and evolutionary classification of lung cancer in never smokers. Nat Genet 53, 1348–1359 (2021).

11. Ding, A., Eisenberg, J. D. & Pandharipande, P. V. The Economic Burden of Incidentally Detected Findings. Radiologic Clinics of North America 49, 257–265 (2011).

12. Novellis, P. et al. Lung cancer screening: who pays? Who receives? The European perspectives. Transl Lung Cancer Res 10, 2395–2406 (2021).

13. Davenport, M. S. Incidental Findings and Low-Value Care. American Journal of Roentgenology 221, 117–123 (2023).

14. Wan, N. et al. Machine learning enables detection of early-stage colorectal cancer by whole-genome sequencing of plasma cell-free DNA. BMC Cancer 19, 832 (2019).

15. Mathios, D. et al. Detection and characterization of lung cancer using cell-free DNA fragmentomes. Nat Commun 12, 5060 (2021).

16. Klein, E. A. et al. Clinical validation of a targeted methylation-based multi-cancer early detection test using an independent validation set. Annals of Oncology 32, 1167–1177 (2021).

17. Fernandez-Uriarte, A., Pons-Belda, O. D. & Diamandis, E. P. Cancer Screening Companies Are Rapidly Proliferating: Are They Ready for Business? Cancer Epidemiology, Biomarkers & Prevention 31, 1146– 1150 (2022).

18. O’Donnell, J. S., Teng, M. W. L. & Smyth, M. J. Cancer immunoediting and resistance to T cell-based immunotherapy. Nat Rev Clin Oncol 16, 151–167 (2019).

19. Patel, A. J. et al. A highly predictive autoantibody-based biomarker panel for prognosis in early-stage NSCLC with potential therapeutic implications. Br J Cancer 126, 238–246 (2022).

20. Yang, R., Han, Y., Yi, W. & Long, Q. Autoantibodies as biomarkers for breast cancer diagnosis and prognosis. Front. Immunol. 13, 1035402 (2022).

21. Lastwika, K. J. et al. Posttranslational modifications induce autoantibodies with risk prediction capability in patients with small cell lung cancer. Sci. Transl. Med. 15, eadd8469 (2023).

22. Schrag, D. et al. Blood-based tests for multicancer early detection (PATHFINDER): a prospective cohort study. The Lancet 402, 1251–1260 (2023).

23. Larman, H. B. et al. Autoantigen discovery with a synthetic human peptidome. Nature Biotechnology 29, 535–541 (2011).

24. O’Donovan, B. et al. High-resolution epitope mapping of anti-Hu and anti-Yo autoimmunity by programmable phage display. Brain Communications 2, fcaa059 (2020).

25. Bodansky, A. et al. Unveiling the proteome-wide autoreactome enables enhanced evaluation of emerging CAR-T therapies in autoimmunity. J Clin Invest (2024) doi:10.1172/JCI180012.

26. Vazquez, S. E. et al. Identification of novel, clinically correlated autoantigens in the monogenic autoimmune syndrome APS1 by proteome-wide PhIP-Seq. eLife 9, e55053 (2020).

27. Mandel-Brehm, C. et al. Kelch-like Protein 11 Antibodies in Seminoma-Associated Paraneoplastic Encephalitis. New England Journal of Medicine 381, 47–54 (2019).

28. Mandel-Brehm, C. et al. Autoantibodies to Perilipin-1 Define a Subset of Acquired Generalized Lipodystrophy. Diabetes 72, 59–70 (2023).

29. Mohan, D. et al. PhIP-Seq characterization of serum antibodies using oligonucleotide-encoded peptidomes. Nature Protocols 13, 1958–1978 (2018).

30. Chen, A., Kammers, K., Larman, H. B., Scharpf, R. B. & Ruczinski, I. Detecting antibody reactivities in Phage ImmunoPrecipitation Sequencing data. BMC Genomics 23, 654 (2022).

31. Raghavan, M. et al. 1 Proteome-wide antigenic profiling in Ugandan cohorts identifies associations between age, 2 exposure intensity, and responses to repeat-containing antigens in Plasmodium falciparum. eLife (2023).

32. Vazquez, S. E. et al. Autoantibody discovery across monogenic, acquired, and COVID-19-associated autoimmunity with scalable PhIP-seq. eLife 11, e78550 (2022).

33. Bulik-Sullivan, B. et al. Deep learning using tumor HLA peptide mass spectrometry datasets improves neoantigen identification. Nature Biotechnology 37, 55–63 (2018).

34. Sidhom, J.-W., Larman, H. B., Pardoll, D. M. & Baras, A. S. DeepTCR is a deep learning framework for revealing sequence concepts within T-cell repertoires. Nat Commun 12, 1605 (2021).

35. Liu, J. et al. A Viral Exposure Signature Defines Early Onset of Hepatocellular Carcinoma. Cell 182, 317–328.e10 (2020).

36. Mann, S. A. et al. Scaled High Throughput Vacuum PhIP Protocol. (2022).

37. Zhao, Y., Tang, H. & Ye, Y. RAPSearch2: a fast and memory-efficient protein similarity search tool for next-generation sequencing data. Bioinformatics 28, 125–126 (2012).

38. Grewal, J. K. et al. Application of a Neural Network Whole Transcriptome–Based Pan-Cancer Method for Diagnosis of Primary and Metastatic Cancers. JAMA Netw Open 2, e192597 (2019).

39. Hosni, M., García-Mateos, G., Carrillo-de-Gea, J. M., Idri, A. & Fernández-Alemán, J. L. A mapping study of ensemble classification methods in lung cancer decision support systems. Med Biol Eng Comput 58, 2177–2193 (2020).

40. Lee, S. H. et al. Multi-view radiomics and dosiomics analysis with machine learning for predicting acute-phase weight loss in lung cancer patients treated with radiotherapy. Phys. Med. Biol. 65, 195015 (2020).

41. Mick, E. et al. Integrated host/microbe metagenomics enables accurate lower respiratory tract infection diagnosis in critically ill children. Journal of Clinical Investigation 133, e165904 (2023).

42. Rackaityte, E. et al. Validation of a murine proteome-wide phage display library for identification of autoantibody specificities. JCI Insight (2023) doi:10.1172/jci.insight.174976.

43. Ma, Q. & He, J. Enhanced expression of queuine tRNA-ribosyltransferase 1 (QTRT1) predicts poor prognosis in lung adenocarcinoma. Ann Transl Med 8, 1658–1658 (2020).

44. Liu, X. et al. PRRC2A Promotes Hepatocellular Carcinoma Progression and Associates with Immune Infiltration. JHC **Volume** 8, 1495–1511 (2021).

45. Bettegowda, C. et al. Detection of Circulating Tumor DNA in Early-and Late-Stage Human Malignancies. Science Translational Medicine 6, 224ra24-224ra24 (2014).

46. Dang, D. K. & Park, B. H. Circulating tumor DNA: current challenges for clinical utility. Journal of Clinical Investigation 132, e154941 (2022).

47. Duffy, M. J. Circulating tumor DNA (ctDNA) as a biomarker for lung cancer: Early detection, monitoring and therapy prediction. TUB 46, S283–S295 (2024).

48. Wagner, S. J. et al. Transformer-based biomarker prediction from colorectal cancer histology: A large-scale multicentric study. Cancer Cell 41, 1650–1661.e4 (2023).

49. Alleman, K. et al. Multimodal Deep Learning-Based Prognostication in Glioma Patients: A Systematic Review. Cancers 15, 545 (2023).

50. Placido, D. et al. A deep learning algorithm to predict risk of pancreatic cancer from disease trajectories. Nat Med 29, 1113–1122 (2023).

51. Jiang, Y. et al. Biology-guided deep learning predicts prognosis and cancer immunotherapy response. Nat Commun 14, 5135 (2023).

52. Suppiah, A. Clinical utility of anti-*p53* auto-antibody: Systematic review and focus on colorectal cancer. WJG 19, 4651 (2013).

53. Steentoft, C. et al. A strategy for generating cancer-specific monoclonal antibodies to aberrant *O* - glycoproteins: identification of a novel dysadherin-Tn antibody. Glycobiology 29, 307–319 (2019).

